# Rapid detection of SARS-CoV-2 infection by multicapillary column coupled ion mobility spectrometry (MCC-IMS) of breath. A proof of concept study

**DOI:** 10.1101/2020.06.30.20143347

**Authors:** Claus Steppert, Isabel Steppert, William Sterlacci, Thomas Bollinger

**Author notes:** Corresponding author: Dr. Claus Steppert, Department of Pulmonology, Thoracic Oncology, Sleep Medicine, Klinikum Bayreuth, Preuschwitzer Str. 101, D-95445 Bayreuth, Germany.

## Abstract

There is an urgent need for screening of patients having a communicable viral disease to cut infection chains.

We could recently demonstrate that MCC-IMS of breath is able to identify Influenza-A infected patients. With decreasing Influenza epidemic and upcoming SARS-CoV-2 infections we went on and also analysed patients with suspected SARS-CoV-2 infections.

75 patients, 34m, 41f, aged 64.4 ± 15.4 years, 14 positive for Influenza-A, 16 positive for SARS-CoV-2, the remaining 44 patients were used as controls. In one patient RT-PCR was highly suspicious of SARS-CoV-2 but initially inconclusive.

Besides RT-PCR analysis of nasopharyngeal swabs all patients underwent MCC-IMS analysis of breath. There was no difference in gender or age according to the groups.

97.3% of the patients could be correctly classified to the respective group by discriminant analysis. Even the inconclusive patient could be mapped to the SARS-CoV-2 group applying the discrimination function.

**Conclusion:** MCC-IMS is able to detect SARS-CoV-2 infection and Influenza-A infection in breath. As this method provides exact, fast non-invasive diagnosis it should be further developed for screening of communicable viral diseases.

Trial registration: ClinicalTrial.gov, NCT04282135 Registered 20 February 2020 - Retrospectively registered, https://clinicaltrials.gov/ct2/show/NCT04282135?term=IMS&draw=2&rank=1

## Introduction

To interrupt infections chains in SARS-CoV-2-disease screening methods are urgently needed. Diagnostic standard is reverse transcription polymerase chain reaction (RT-PCR) but deep nasopharyngeal swabs taken by trained personal are required and rapid PCR-techniques still take more than 30 minutes.^1^ Especially for screening at airports or other sites where rapid screening of asymptomatic patients is demanded this method is logistically challenging and takes usually by far longer than 30 min. Additionally especially in the scope of developing countries RT-PCR is far too expensive. So, there is still a need for a faster, really noninvasive screening tool.^2^ A screening tool that can be easily used and prevents false negative results would fulfil this demand.

In classic antiquity without other diagnostic tools physicians had to rely on their basic senses, seeing, touching, hearing and smelling.^3^ With improved technical possibilities these skills have been moved in the background. Only few scents physicians are taught in medical school like acetone for ketoacidosis and ammonia for liver disease. It is well known that different bacteria smell differently. *Pseudomonas aeruginosa* have a fruity scent while *Escherichia coli* smells faecal. Based on their better olfactory senses animals have been trained to smell infectious diseases but lack of reproducibility precludes wider application.^4^

The scents of infectious diseases are volatile organic compounds (VOC) that are released by the metabolism of the germ or the host. There are different technical approaches to discriminate pathogens or diseases based on VOCs but none has been used regularly in clinical practice.^6^

Gas chromatography coupled ion mobility spectrometry (GC-IMS) has proven to discriminate bacterial infections in vitro as in breath.^6–9^

Compared to bacteria viruses have no own metabolism. So using scents can only rely to the host response to the viral infection.^10^ Currently there are only few studies addressing this issue. Gas chromatography coupled mass spectrometry (GC-MS) was able to detect Influenza infection of cell cultures in vitro.^11^

As GC-MS is not feasible for point-of-care diagnostics there are attempts to train dogs to sniff for viral diseases, currently for SARS-CoV-2 associated disease.^12,13^ In a recently published study dogs were trained to smell samples from COVID-19 patients with an average detection rate of 94%.^14^

In a recent study we could demonstrate that Influenza-A infection can also be detected in the breath of Influenza-A infected patients by multicapillary-column-coupled ion mobility spectrometry (MCC-IMS).^15^ Therefore, we extended this study to analyse whether MCC-IMS is also able to detect SARS-CoV-2- infection in breath.

## Methods

During the influenza-A epidemic 2020^15^ 11 male, 13 female, aged 66 ± 14.2 years and during the SARS-CoV-2-pandemic 23 male, 28 female, aged 63 ± 16 years with suspected infections were asked to participate in the study. The study was approved by the Ethics Committee of Erlangen University #426_18_B and registered at ClinicalTrials.gov (NCT04282135).

After routine RT-PCR analysis of nasopharyngeal swabs 14 of the patient were positive for influenza-A, 16 were positive for SARS-CoV-2, the remaining 44 patients were used as controls.

Breath samples were taken and analysed by MCC-IMS. Patients were recruited between April 8^th^ 2020 and May 7^th^ 2020. Unfortunately, many patients could not be included as they were too sick and had to be transferred to the intensive care unit or were unable to provide written consent. As the time went on, measures of social distancing and segregation were successful leading to decreasing numbers of SARS-CoV-2 patients.

### PCR

SARS-CoV-2 was tested by taking a deep nasopharyngeal swab applying the “Xpert^®^ Nasopharyngeal Sample Collection Kit for Viruses” (Cepheid, Maurens-Scopont, France) and performing real time PCR by applying the Allplex 2019-nCoV Assay (Seegene, Seoul, South Korea) on the CFX 96 Real-Time Sytem (BioRad, Feldkirchen, Germany) after extracting RNA by using the StarMag 96 UniTube Kit (Seegene) on the SGPrep32 extraction system (Seegene). Due to shortage in supply RNA was in part alternatively extracted applying the QiaAmp DSP Virus Spin Kit (Qiagen, Hilden, Germany) using the Qiacube automated extraction system (Qiagen). Influenza PCR was performed with the “Xpert® Xpress Flu/RSV” (Cepheid) on the “Infinity” (Cepheid).

### Breath-Sampling and MCC-IMS

For the ion mobility spectrometry we used the MCC-IMS-device from STEP Sensortechnik und Elektronik, Pockau, Germany (STEP IMS NOO). The device is distributed as a medical device (In-Vitro-Diagnostic) in combination with an evaluation Software as “MultiMarkerMonitor^®^” by Graupner medical solutions GmbH, Geyer, Germany.

All patients were connected by a foam cuffed oxygen catheter (#01442958, Asid-Bonz, Herrenberg, Germany) via a 0.22μm Filter (Navigator Lab Instruments, Tinajin, China) and a Perfusor Line (B Braun, Melsungen, Germany) directly to the MCC-IMS.

Patients were instructed to take a deep breath and to exhale slowly through the nose. During the exhalation breath was sampled for 10s.

The STEP device directly draws the sample by an internal pump (200mL/min) into the analysing circuit without any pre-analytical procedures.

In an inert sampling loop of 2 mL the sample is standardized for volume. Then the sample is pre-separated by isothermally heated multicapillary gas chromatography column (60°C) into single analytes, which enter the IMS unit based on their retention times. In the IMS unit the analytes are ionized by beta radiation of a tritium source below the free limit for radiation (99 MBq). Afterwards the generated ions are accelerated in a 50-mm-long drift-tube under the influence of an electric field (400 V/cm) towards the detector which is also tempered to 60°C. On their way the positive ions collide with air molecules from the drift gas (400 mL/min) flowing in the opposite direction and are separated depending on their ion mobilities and detected by the collector electrode sampled every 10μs. The received IMS spectra are stored internally in the device and later analysed offline.

The used IMS device is equipped with a circulation filter and internal gas circulation. Using a circulation pump, ambient air filtered by an activated carbon filter was provided as drift gas and analysis gas (20 mL/min) to the device. Compared to other IMS devices there is no need for a special analysis gas.

### Data analysis

The VOCs are characterized by their retention time in the MCC and the drift time in the IMS. One spectrum over 2048 measurement points every 10 μs (in total 20.48ms) is obtained every second for a total time of 240s.

These spectra can be visualized on a heatmap with retention time on the Y-axis and the drift time on the X-axis.

To decrease the complexity of the data we used a proprietary cluster analysis software using support vector machine (European Patent EP 2 729 801 B1).^16^ After baseline correction for noise the software determines the clusters based on the signal threshold and categorizes them by retention time and drift time. Depending on these parameters the clusters are numbered assuming that every cluster represents a distinct VOC.

### Statistical Analysis

Due to the small sample size and the lack of normal distribution Mann-Whitney-White U-Test and Kruskal-Wallis Test were applied for differences in patient characteristics.

Patients from both sub studies where neither Influenza-A nor SARS-CoV-2 was found in the PCR were combined as controls for the combined dataset.

To exclude cross-correlated clusters we performed a stepwise canonical discriminant analysis for optimal minimization of Wilks Lambda. For entering or removing variables from the model F significances of 0.05 and 0.1 were used.

For the statistical analysis we used IBM SPSS 22.0 (IBM, Armonk, NY).

### Results

Age and infection markers as well as biometrics were not different between the groups. The only difference was the reduced leukocyte count in the SARS-CoV-2-positive population (Tab. 1). The influenza-A positive patients were included to verify that the identified specific clusters of SARS-CoV-2 not simply reflect metabolic changes of viral infection.

**Table 1.**
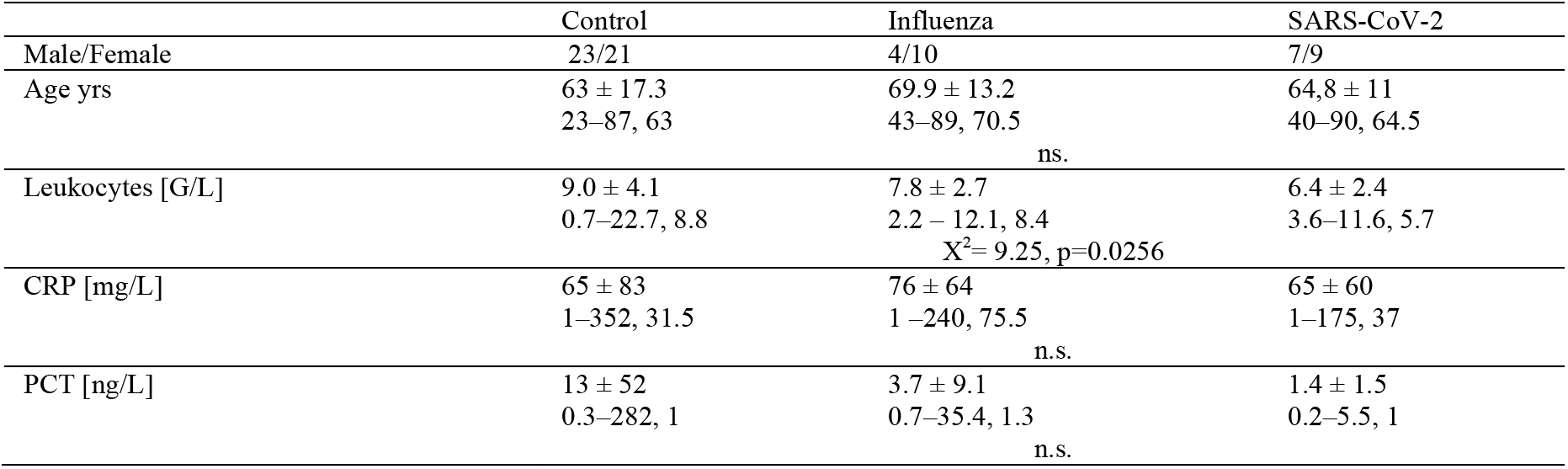
Patient characteristics for the combined dataset. For the continuously measurable parameters in the first row mean and SD, in the second row range and median are reported. n.s. = not significant.

Though the patients were triaged as suspected by the symptoms at presentation there was significant more fever, cough and dyspnea in the SARS-CoV-2- positive group as well as in the Influenza group. Only gastrointestinal symptoms did not differ between the groups. Compared to Influenza dyspnea was significantly more common in the SARS-CoV-2 group. (Fig. 1).

**Figure 1.**
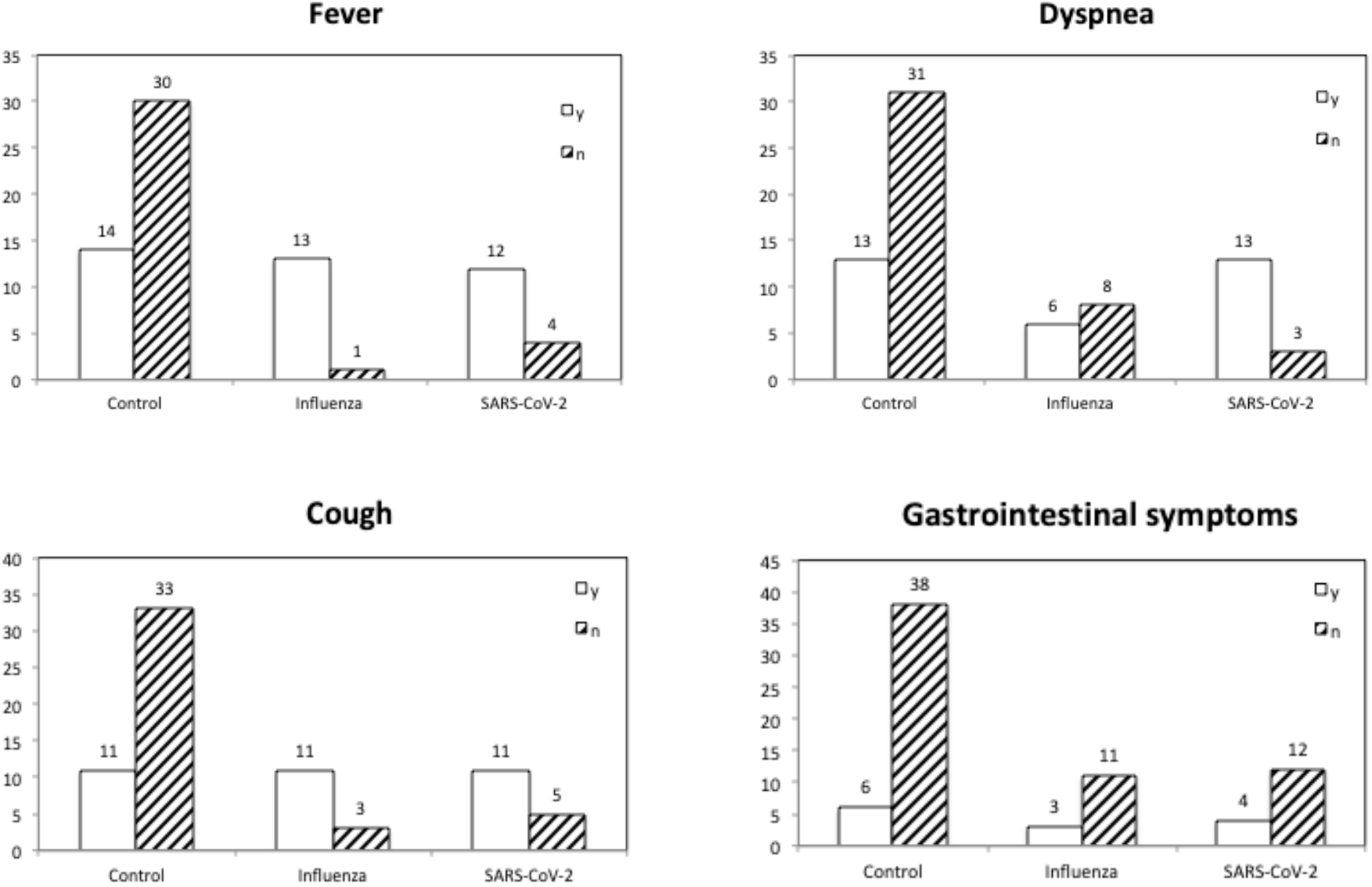
Symptoms at admission.

**Table 2.** Patient symptoms. n.s. = not significant.

|  | Control | SARS-CoV-2 | Influenza A |
| --- | --- | --- | --- |
| Fever | 14/44 | 12/16 | 13/14 |
| | | $X^2=20.86, p<0.001$ | |
| Dyspnea | 13/44 | 13/16 | 6/14 |
| | | $X^2=12.78, p=0.001$ | |
| | | $X^2=12.78, p<0.0001$ | |
| Cough | 11/44 | 11/16 | 11/14 |
| | | $X^2=17.18, p<0.001$ | |
| Gastrointestinal Symptoms | 6/44 | 4/16 | 3/14 |
|  |  | n.s. |  |
| Pathological X-Ray | 10/41 | 14/16 | 3/14 |
| | | $X^2=13.27, p<0.001$ | |
|  |  | n.s. |  |
| | | $X^2=21.49, p<0.001$ | |

In one patient SARS-CoV-2- PCR was initially inconclusive as the manufacturer (Seegene) meanwhile redefined the definition for positive test results this patient would now be categorized as positive. This result was reproducible in a second test.

155 clusters were found that were further used for the multivariate analysis. By two canonical discrimination functions 97.3% of the cases were correctly classified at cross-validation, even between influenza-A and SARS-CoV-2.

At cross-validation there was only one control misclassified as influenza-A and one as SARS-CoV-2, respectively. There were no false negatives in this analysis.

**Table 3.**
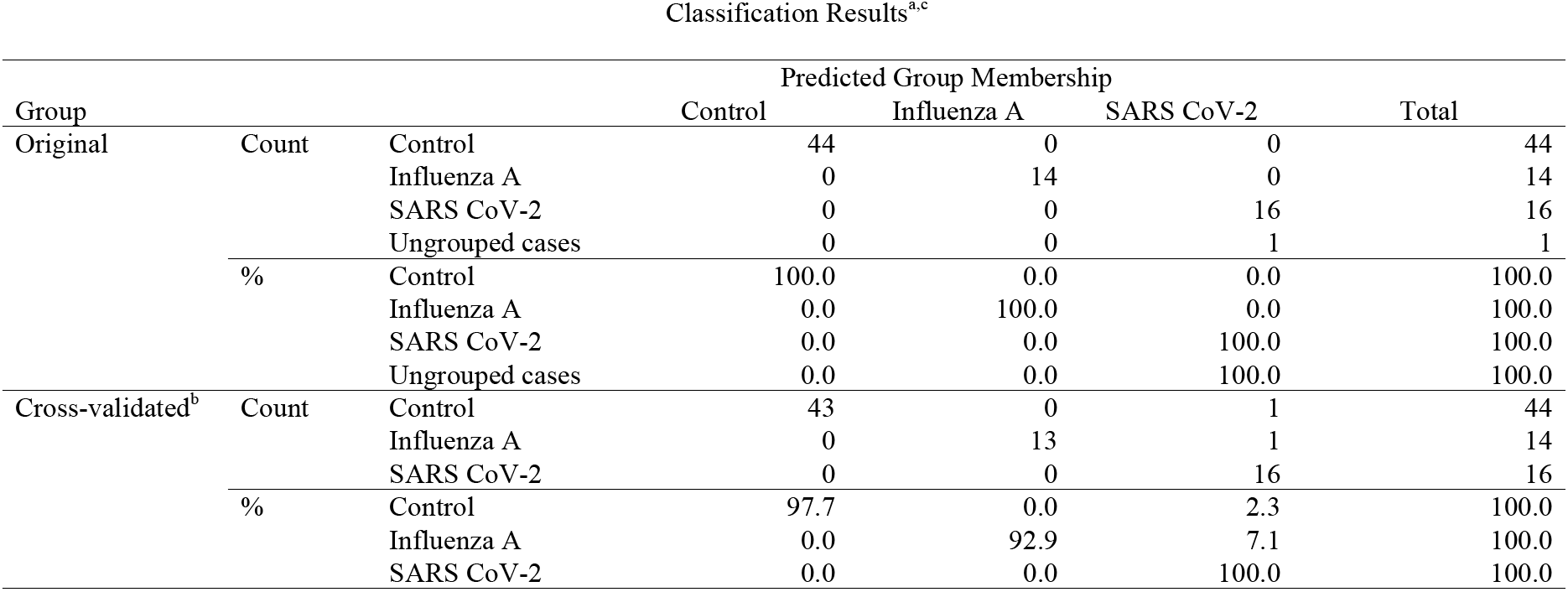
Classification results for the canonical discriminant analysis a. 100.0% of original grouped cases correctly classified. b. Cross validation is done only for those cases in the analysis. In cross validation. each case is classified by the functions derived from all cases other than that case. c. 97.3% of cross-validated grouped cases correctly classified.

Discriminant analysis is able to explain 100% of the variance. In the scatter plot of the two discriminant functions all 3 groups are separated nicely (Fig. 2). The patient with the initial inconclusive PCR result had low viral load and is never the less mapped to the SARS-CoV-2 group. For this patient one RT-PCR showed only a signal of the *E*- as well as *N*-gene with PCR crossing point of 35.44 for both genes. A second PCR performed two days later gave a positive signal of the *RdRP*-gene with a crossing point of 37.65 while the *E*- and *N*-gene were then negative.

**Figure 2.**
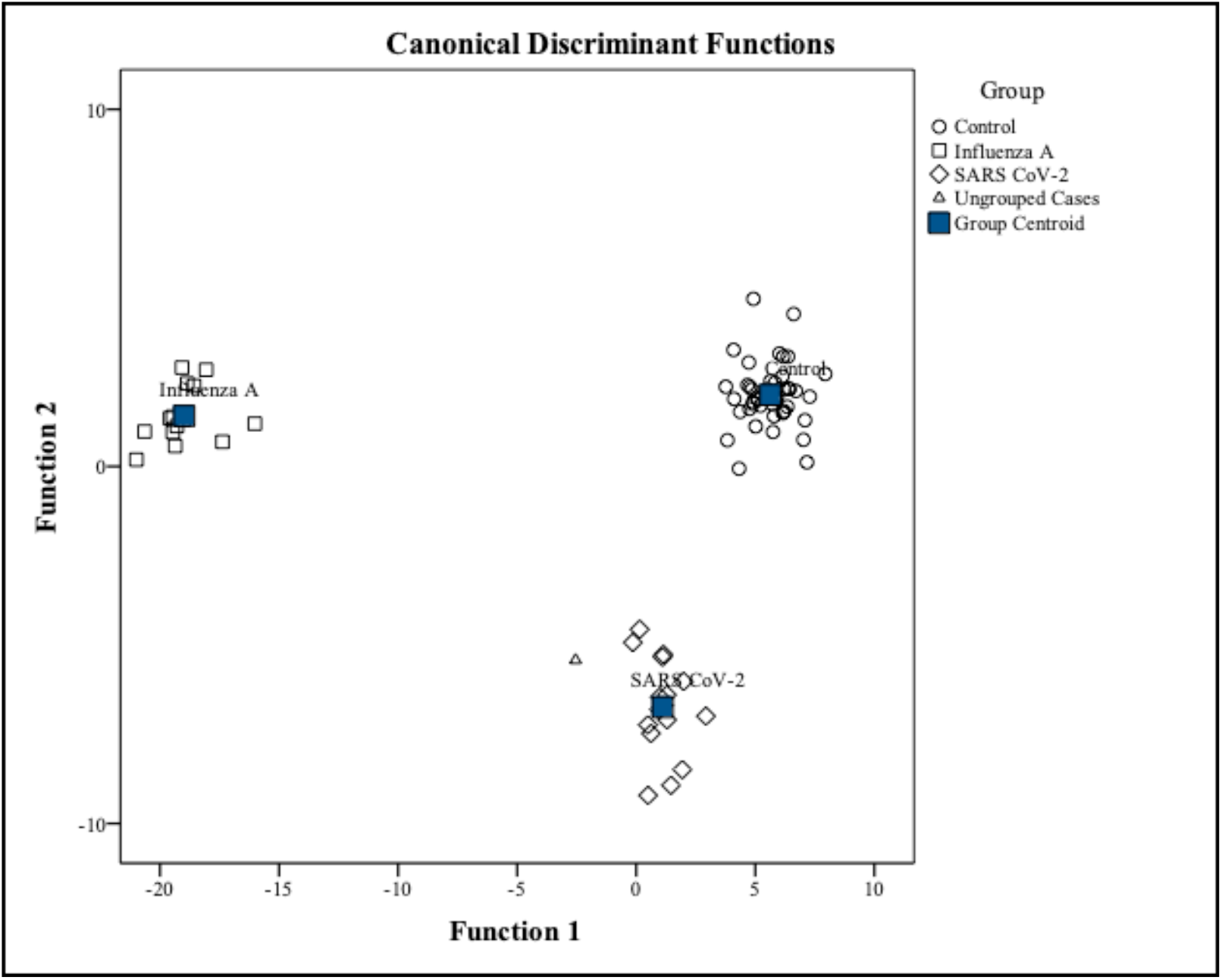
Results of the canonical discrimination analysis. Controls as circles, Influenza-A as squares, SARS-CoV-2 as diamonds and the only unclassified as a triangle. Note, that later on the unclassified patient was re-classified as SARS-CoV-2 positive due to changes of the SARS-CoV-2 PCR interpretation criteria by the manufacturer.

## Discussion

Today RT-PCR is considered the gold standard in the diagnosis of SARS-CoV-2- infection but even in trained hands a false negative rate of about 25% and a false positive rate of 2.3-6.9% has to be expected.^17^ However, false negative results are often caused by low swab and sampling technique quality while in here only well trained staff performed the sampling applying brushed swab from Copan which can be considered as high quality. Hence, we did only notice false negative nasopharyngeal swabs in patients who were either in the pneumonia phase of COVID-19 or were at the detection limit.

Our unpublished experience was that patient at the detection limit usually are at the very end of a COVID-19 and are most likely not infective anymore.^18^

Currently SARS-CoV-2 infections are rising in several countries causing tremendous cost. In underdeveloped countries neither trained staff nor financial power to afford RT-PCR for mass screening is available.^2^

To the best of our knowledge our study is the first showing that breath-analysis is able to discriminate SARS-CoV-2 infected patients from controls with respiratory infections and Influenza infection as well. Even one suspected but not clearly PCR-confirmed patient could be assigned to the SARS-CoV-2 group and turned out to be positive later.

We used MCC-IMS because of the ease of application. The STEP-IMS device does not need any pre-analytic procedures or test gases. So, no shortage of swabs, tubes or reagents has to be faced in the scope of a pandemic.

The device draws the breath into the system by an internal pump. This simplifies the sampling compared to other IMS devices where absorption/desorption tubes are needed.^9^

The only task to be fulfilled by less trained staff is to introduce the foam-cuffed catheter into a nostril of a spontaneously breathing individual and to hit a key to start the measurement.

For our study written consent of the patient was a prerequisite. Therefore, we had to exclude demented, delirious or too severely ill patients not able to consent but this should be no constraint for the method in real life.

It could be argued that IMS only provides peaks according to retention time and drift time while mass spectrometry (MS) is able to chemically describe the VOCs but also in MS not all peaks are clearly assigned to a chemical substance and are therefore also only numbered or characterised by the time of flight.^6^ Furthermore, even though it is academically interesting to identify the relevant peaks in the breath of COVID-19 patients it is dispensable for SARS-CoV-2 screening. Hence, we think that not knowing exactly the chemical structure of the VOC is no detriment as another attempt to screen for SARS-CoV-2 is by the smell of trained dogs.^13,14^ Like scent dogs, a fingerprint of peaks should enable the classification of the odour of infected patients.

As the scent of the breath does not rely on the virus itself but on the host response to the infection cross-reactivity of breath analysis with other viral infections has to be expected.^10^ As MCC-IMS could differentiate between SARS-CoV-2 and Influenza-A infection we assume that different viruses cause at least to a certain extend different host responses and therefore produce different fingerprints of IMS spectra. However, this needs to be addressed in future studies.

Similar to antibody tests an overlap of the VOCs with other corona virus infections has to be anticipated. But this is a constraint every analysis of metabolomics has to face. However, within the current pandemic we detected almost no endemic corona viruses in our adult patients.

Compared to other breath analysis studies we did not require fasting before the sampling. Though fasting state may reduce interferences with other metabolism it will not be feasible for large scale screening.

Another necessity for a further progress of breath analysis in screening for infections is the extension of the study to other ethnicities and civilisations to investigate whether ethnos and life style needs to be considered for the analysis.

One drawback of our study is the limited number of patients. As pointed out already many patients with SARS-CoV-2 were not able to give informed consent. Another point is the weakening of the SARS-CoV-2 wave in Germany end of April as this led to a slowing of accrual.

We therefore assess this study as a proof-of-concept and encourage other researches to further investigate breath analysis by MCC-IMS for the detection of SARS-CoV-2 infections. We are currently developing a point-of-care prototype with an instant analysis of the data as this will be the relevant step for large scale screening.

As MCC-IMS is fast, non-invasive and does not need any reagents or pre-analytical procedures it seems promising for a screening device even in underdeveloped countries or air travel. In conclusion we identified a quick and cheap way of large-scale SARS-CoV-2 testing.

## Conclusion

Breath analysis using MCC-IMS is able to discriminate between Influenza-A, SARS-CoV-2- infections and - controls in a few minutes.

As this method is completely non-invasive and does not need any reagents or pre-analytic procedures it seems promising for a mass screening device even in underdeveloped countries. We encourage further trials to use this technique in different patient settings.

### Declarations

The study was approved by the ethics committee of Erlangen University #426_18_B

All patients gave informed written consult

There was no external funding

### Author’s Contribution

Claus Steppert: Literature search, study design, data collection, data analysis, data interpretation, writing, review

Isabel Steppert: Data analysis, writing, review

William Sterlacci: Review

Thomas Bollinger: Data analysis, writing, review

All authors have consented and approved the submission

The authors state that there is no conflict of interests.

Data used for statistical calculations as well as IMS settings and raw output of discriminant analysis is publicly available as online supplement. Raw Data (~1GB) is available on request.

## Data Availability

Data not publicly available, available by authors on request

## Acknowledgement

The authors thank Dr. G. Becher and Graupner medical solutions GmbH for providing the MCC-IMS device, the cluster analysis software and especially Dr. Becher for his support in analysing the MCC-IMS data.

